# X-Chromosome-Wide Association Study Identifies Novel Genetic Signals for Heart Failure and Subtypes

**DOI:** 10.64898/2026.04.21.26351435

**Authors:** Junling Ren, Chang Liu, Qin Hui, Maryam Rahafrooz, Nicole M Kosik, Kevin Urak, Jennifer Moser, Sumitra Muralidhar, Alexandre C Pereira, Kelly Cho, J Michael Gaziano, Peter W F Wilson, VA Million Veteran Program, Lawrence S Phillips, Yan V Sun, Jacob Joseph

**Author notes:** Equal contribution (Junling Ren, Chang Liu). **Corresponding Author:** Jacob Joseph, MD Cardiology Section, VA Providence Healthcare System, Providence, Rhode Island, USA.

## Abstract

**Background:** Heart failure (HF) is a major and growing public health problem, and prior studies support a meaningful genetic contribution to HF susceptibility. Clinically, HF is commonly categorized into the major clinical sub-types of HF with reduced ejection fraction (HFrEF) and HF with preserved ejection fraction (HFpEF), which differ in pathophysiology and clinical profiles. However, previous genome-wide association studies have focused on autosomal variation and have routinely excluded the X chromosome, leaving X-linked genetic contributions to HF and its subtypes under-characterized.

**Methods:** We performed X-chromosome wide association study (XWAS) utilizing directly genotyped data from 590,568 Million Veteran Program participants, including 90,694 HF cases across European, African, Hispanic, and Asian Americans. Sex- and ancestry-stratified logistic regression was used with XWAS quality control measures, adjusting for age and population structure, followed by fixed-effects multi-ancestry meta-analysis. Functional annotation, gene-based testing, fine-mapping, and colocalization were performed. We replicated genetic associations with all-cause HF in the UK Biobank.

**Results:** In the multi-ancestry meta-analysis, we identified five X-chromosome-wide significant loci for all-cause HF, five for HFrEF, and one locus for HFpEF in males. No loci reached significance in female-specific analyses. In sex-combined analyses, we identified six loci for all-cause HF and four for HFrEF. The strongest and most emphasized signals mapped to genes were *BRWD3, FHL1*, and *CHRDL1*. Ancestry-specific analyses revealed additional loci, including *NDP* and *WDR44* in African ancestry and *PHF8* in Hispanic ancestry. One locus, *BRWD3*, was replicated in UK Biobank HF cohort. Integrated post-GWAS analyses (fine-mapping, colocalization and pleiotropy trait association studies) reinforced the biological plausibility of the X-linked signals.

**Conclusions:** This multi-ancestry, sex-stratified XWAS identifies X-linked genetic contributions to HF and its subtypes and highlights the role of X-chromosome in heart failure pathogenesis.

## Introduction

Heart failure (HF) is a major and growing public health problem, associated with substantial morbidity, mortality, and health-care utilization. HF is affecting approximately 7 million Americans, a number projected to increase to 8.5 million by 2030 [1]. HF is clinically categorized by a single measure, left ventricular ejection fraction into the major clinical sub-types of HF with reduced ejection fraction (HFrEF) and HF with preserved ejection fraction (HFpEF), which differ in underlying pathophysiology and patient profiles [2, 3]. Notably, marked sex differences are observed across the HF spectrum: men are more likely to present with HFrEF, predominantly linked to ischemic loss and myocardial fibrosis, whereas women predominate in HFpEF, primarily driven by microvascular dysfunction and inflammatory pathways [4-6].

A substantial genetic component contributes to HF susceptibility, with an estimate of 26-34% from a Swedish study [7]. A multi-ancestry HF genome-wide association studies (GWAS) meta-analysis reported 47 risk loci [8]. Subtype-focused analyses have demonstrated distinct genetic architectures for HFrEF versus HFpEF [9], and a recent large-scale GWAS meta-analysis further expanded locus discovery and provided etiologic insights across HF subtypes [10]. Despite this progress, most HF GWAS discovery has emphasized autosomes, leaving the contribution of the X chromosome under-characterized.

The X chromosome is biologically and analytically distinct from autosomes [11, 12] and challenges in analysis have led to routine exclusion or inconsistent handling of X-chromosomal variation in GWAS. Recent methodological advances have enabled rigorous X-chromosome-wide association studies (XWAS) at scale [11]. Because X-chromosomal variation is intrinsically tied to sex-specific biology, we report the first large-scale sex-stratified XWAS of HF and its subtypes in a multi-ancestry cohort. This study aims to prioritize genetic loci that could inform the development of targeted therapies and interventions to prevent or mitigate HF risk.

## Methods

### Discovery Cohort: The Million Veteran Program (MVP)

We conducted analyses in the MVP, an ongoing, national research cohort embedded within the U.S. Department of Veterans Affairs (VA) health care system [13]. MVP enrolls Veterans receiving care at VA facilities nationwide. Participants provided written informed consent to donate a blood specimen and to link survey responses with longitudinal VA electronic health records (EHR). Data available for research include baseline and lifestyle surveys, biospecimens, and comprehensive EHR domains including administrative codes, clinical notes, medication prescriptions, laboratory values, and imaging reports. Recruitment began in 2011, and the cohort has grown to >1,000,000 participants, with broad ancestry representation that supports multi-ancestry genetic studies. Genetic ancestry was assigned using the harmonized ancestry and race/ethnicity (HARE) approach [14]. Briefly, we computed genetic principal components (PCs) from linkage-disequilibrium-pruned autosomal variants, then applied a supervised machine-learning model that integrates these genetic PCs with self-identified race/ethnicity from VA surveys/EHR to produce mutually exclusive ancestry groups, including European, African, Hispanic, and Asian.

### HF Phenotyping in the MVP

We identified HF cases in the MVP by the presence of at least one inpatient or outpatient diagnosis code for HF (ICD-9: 428.x; ICD-10: I50.x) and required an echocardiogram performed within 6 months of the index HF diagnosis [9]. Left ventricular ejection fraction (LVEF) values were ascertained using a validated VA natural-language-processing (NLP) pipeline applied to echocardiography reports. To maximize capture (including studies performed outside VA but documented in the record), the NLP pipeline also parsed nuclear medicine reports, cardiac catheterization reports, history and physicals, progress notes, discharge summaries, and cardiology notes, with accuracy confirmed by blinded physician review[15]. HF was subtyped as HFrEF when LVEF ≤ 40% and HFpEF when LVEF ≥ 50% using the LVEF closest to the diagnosis. Controls were MVP participants without any HF diagnosis codes.

### X-Chromosome Data Quality Control in the MVP

Genotyping was performed using Affymetrix Axiom Biobank Arrays. An overview of the workflow is shown in **Figure 1**. Briefly, we first applied standard, array-based quality control (QC) procedures to the autosomal data, including filters on sample and variant call rate, Hardy-Weinberg equilibrium (HWE), heterozygosity outliers, and relatedness. Related individuals (second-to third-degree relatives or closer) were identified using KING [16], and one individual from each related pair was excluded. After completion of general QC, X-chromosome genotypes were extracted and the pseudoautosomal regions (PARs) were removed; approximately 13,000 non-PAR X-chromosome variants remained at this stage.

**Figure 1.**
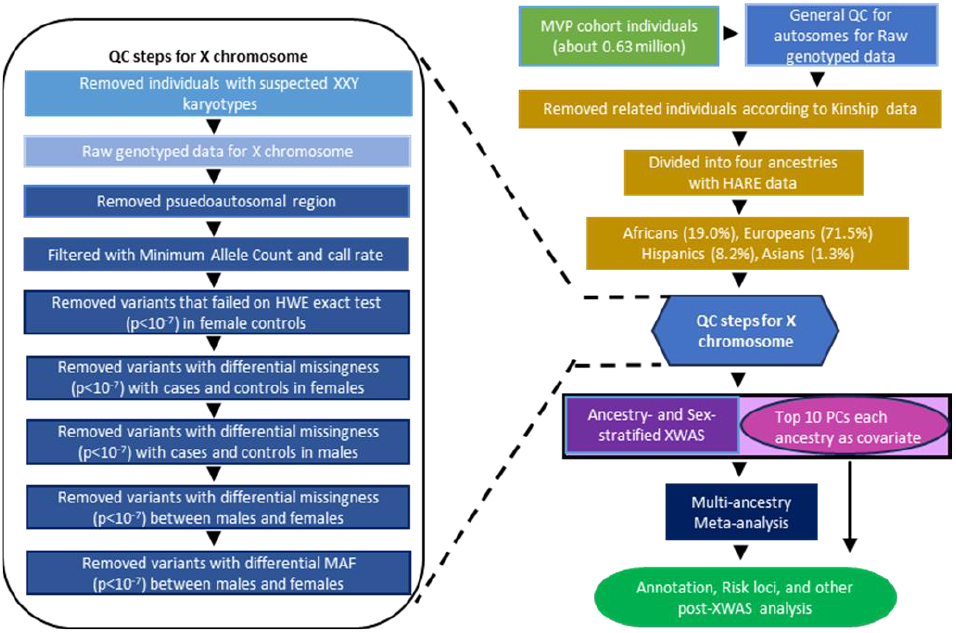
Schematic diagram datasets and XWAS workflow. MVP: Million Veteran Affair; PCs: Principal Components; XWAS: X chromosome -wide association studies; QC: Quality Control; HARE: Harmonized Ancestry and Race/Ethnicity, HWE: Hardy-Weinberg Equilibrium; MAF: Minor Allele Frequency.

After assigning participants to four ancestry groups using HARE, we then implemented an X-chromosome-specific QC pipeline to account for hemizygosity in males and sex-specific genotyping artifacts. Briefly, we restricted analyses to participants who passed autosomal QC and applied additional sample-level checks to confirm genetic sex and exclude sex chromosome aneuploidies. Sex discrepancies were evaluated by comparing self-reported sex with genetically inferred sex. Individuals with suspected abnormal karyotypes (e.g., XXY or XYY) were identified using the distribution/median of Log R Ratio (LRR) intensities on X and Y and excluded. At the variant level, we applied probe-sequence/array-specific QC filters and removed Single-nucleotide polymorphisms (SNPs) with call rate <95% or low allele counts. We excluded SNPs departing from HWE in female controls (P < 1×10^-7^) and removed SNPs showing differential missingness (P < 1×10^-7^) between cases and controls within females, within males, or between sexes. Finally, SNPs with significantly different minor allele frequencies (MAFs) between males and females (P < 1×10^-7^) were excluded. Sex differences in allele frequency were evaluated using sdMAF [17]. After all X-chromosome QC steps, approximately 11,000 high-quality, non-PAR genotyped SNPs remained for association analyses.

### XWAS in the MVP

To minimize the multiple-testing burden, the primary discovery analysis was conducted using directly genotyped data. Association analyses were conducted using XWAS 3.0 [18]. For all-cause HF, HFrEF and HFpEF, analyses were stratified by sex and by genetic ancestries, including European, African, Hispanic and Asian. Within each ancestry-sex stratum, logistic regression model was used to estimate the association with HF and its subtypes, adjusting for age at enrollment and top 10 autosomal PCs, since population structure is best captured by autosomes and avoids confounding PCs with sex or X chromosome’s unique inheritance (e.g., sex-specific ploidy, X-inactivation, pseudoautosomal regions or sex-biased admixture) [19]. This strategy follows established practices in XWAS studies and ensures that ancestry correction reflects broad genomic background rather than X-specific artifacts. The meta-analysis integrating four genetic ancestries was conducted using the GWAMA (Genome-Wide Association Meta-Analysis) software, which incorporates fixed effect models [20].

### X-Chromosome-Wide Significance Threshold

The conventional autosomal genome-wide threshold of p < 5×10^-8^ was not applied, given that the association tests were restricted to the single X-chromosome with fewer independent tests on 11,000 variants. To determine an appropriate significance cutoff, Bonferroni corrected p value was calculated by dividing 0.05 by the number of effective tests (N_eff_), as described in prior studies[21, 22]. Briefly, N_eff_ was calculated using the equation below, where N is the number of variants, L is the R^2^ correlation coefficient between all variants (v) present in datasets (L is a matrix with the size of v by v), i and j are matrix indexes[23]. The effective number of tests and final X-chromosome-wide significance cutoffs for each ancestry-sex stratum are summarized in **Table S1**.

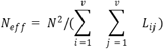

### Functional Annotation, Credible Set Analysis and Colocalization

All XWAS results were functionally annotated using FUMA [24]. SNPs with p values less than the Bonferroni corrected p value threshold and were pairwise independent, i.e., linkage disequilibrium (LD) r^2^<0.6, were kept as independent significant SNPs. From these, sentinel SNPs were selected at r^2^<0.1 using ancestry-matched LD. Genomic risk loci were created by merging LD blocks whose lead signals were within 250 kb. For variants annotated with multiple nearby genes, the closest gene was selected. In cases where the nearest gene was a noncoding RNA, the nearest protein coding gene was reported. Gene-based association analyses were performed using MAGMA [25] to assess gene-level associations.

To pinpoint potential causal variants associated with HF and subtypes, we conducted credible set analyses integrating LD information and Bayesian methods. We first used the LDlinkR R package [26] to obtain variants in LD with each significant lead SNP, using the 1000 Genomes Project as the reference panel [27]. These LD-informed variant groups were used as inputs for downstream fine-mapping. Next, we applied the corrcoverage R package [28] to perform Bayesian fine-mapping, generating credible sets of variants most likely to harbor the causal signal. Credible sets were defined using posterior inclusion probability (PIP) thresholds of 0.90 and 0.95 to represent high-confidence levels.

To assess potential shared genetic regulation, colocalization analysis was performed using the *coloc* R package [29]. For each lead SNP, we defined the locus as ±250 kb and extracted all variants within this window. We estimated posterior probabilities for the five competing hypotheses: PPH0, no association with either trait in the region; PPH1, association with the all-cause HF/HFrEF/HFpEF XWAS trait only; PPH2, association with the expression quantitative trait locus (eQTL) trait only; PPH3, both traits are associated but driven by distinct causal variants; and PPH4, both traits are associated and share a single causal variant (evidence of colocalization). Regional GWAS-eQTL colocalization plots were generated using the locuscomparer package [30].

To investigate broader phenotypic associations of the XWAS risk loci, we queried lead variants from the multi ancestry and ancestry stratified XWAS in the UK Biobank PheWeb resource (https://pheweb.org/UKB-TOPMed/) and summarized associations with p < 1E^-5^.

### Replication of All-cause HF Loci in the UK Biobank

To independently replicate the X-chromosome association signals identified in MVP, we performed association analysis of all-cause HF in the UK Biobank (UKB), a large prospective population-based cohort of approximately 500,000 participants recruited across the United Kingdom. UKB participants were genotyped on the UK Biobank Axiom array, and genotype data were centrally processed and imputed by UKB using reference panels described previously [31]. All-cause HF cases and controls were ascertained from linked longitudinal health outcomes through hospital inpatient records, primary care, death registry and self-reported conditions linked to hospital records. HF cases were defined using ICD-10 diagnosis codes of I50. Controls were participants without HF diagnosis codes. Participants of European ancestry were included, and related individuals (up to approximately third-degree relatives) were excluded.

For replication testing in the UKB, lead SNPs from the MVP discovery analysis were extracted from UKB genotype data and harmonized to the same effect allele to ensure consistent direction-of-effect comparison. Association testing was performed using PLINK [32] and logistic regression. Models were adjusted for age and the top 10 autosomal PCs. The sex-combined model additionally adjusted for sex. A lead SNP was considered successfully replicated if it showed p < 0.05 in UKB and demonstrated a consistent direction of effect relative to MVP.

## Results

The MVP analytic sample included 590,568 participants (90,694 HF cases and 499,874 controls and was predominantly male (537,457 men; 53,111 women). Among men, there were 87,390 HF cases (including 41,515 HFrEF and 37,397 HFpEF) and 450,067 controls; among women, there were 3,304 HF cases (including 1,154 HFrEF and 1,944 HFpEF) and 49,807 controls. HF cases were older than controls (men: 68.18 vs 61.37 years; women: 61.63 vs 49.44 years) and had higher BMI compared to controls in both sexes. The overall cohort comprised 71.96% European, 18.56% African, 8.13% Hispanic, and 1.32% Asian ancestry participants. Cardiometabolic comorbidities were substantially more prevalent among HF cases than controls, including diabetes, chronic kidney disease, hypertension, coronary artery disease, peripheral vascular disease, and atrial fibrillation, **Table 1**. The replication analysis dataset of the UKB included 14,794 HF cases and 322,000 controls in the sex-combined analysis. We additionally conducted sex-stratified analyses including 9,559 cases and 146,401 controls in males, and 5,235 cases and 175,599 controls in females, **Table S2**.

**Table 1.** Cohort Characteristics of the Million Veteran Program.

| Characteristic | Male |  |  | Female |  |  | Overall Cohort |  |  |
| --- | --- | --- | --- | --- | --- | --- | --- | --- | --- |
|  | HF Cases<br>N=87,390<br>N<sub>HEpFF</sub> =37,397<br>N<sub>HFrEF</sub> =41,515 | Controls<br>N=450,067 | ALL<br>N=537,457 | HF Cases<br>N=3,304<br>N<sub>HEpFF</sub> = 1,944<br>N<sub>HFrEF</sub> = 1,154 | Controls<br>N= 49,807 | ALL<br>N=53,111 | HF Cases<br>N = 90,694<br>N<sub>HEpFF</sub> = 39,341<br>N<sub>HFrEF</sub> = 42,669 | Controls<br>N = 499,874 | All<br>N=590,568 |
| Age at Enrollment (years) | 68.18 (0.03) | 61.37 (0.02) | 62.48 (0.02) | 61.63 (0.21) | 49.44 (0.06) | 50.20 (0.06) | 67.94 (0.03) | 60.18 (0.02) | 61.37 (0.02) |
| European (%) | 65,349 (74.78) | 327,757 (72.82) | 393,106 (73.14) | 2,226 (67.37) | 29,656 (59.54) | 31,882 (60.03) | 67,575 (74.51) | 357,413 (71.51) | 424,988 (71.96) |
| African (%) | 16,724 (19.14) | 77,451 (17.21) | 94,175 (17.52) | 940 (28.45) | 14,494 (29.10) | 15,434 (29.06) | 17,664 (19.48) | 91,945 (18.39) | 109,609 (18.56) |
| Hispanic (%) | 4,809 (5.50) | 38,366 (8.52) | 43,175 (8.03) | 120 (3.63) | 4,729 (9.39) | 4,849 (9.13) | 4,929 (5.43) | 43,095 (8.62) | 48,024 (8.13) |
| Asian (%) | 508 (0.58) | 6,493 (1.44) | 7,001 (1.30) | 18 (0.54) | 928 (1.86) | 946 (1.78) | 526 (0.58) | 7,421 (1.48) | 7,947 (1.32) |
| BMI (kg/m<sup>2</sup>) | 30.90 (0.02) | 29.39 (0.01) | 29.63 (0.01) | 32.21 (0.13) | 29.86 (0.03) | 30.00 (0.03) | 30.94 (0.02) | 29.43 (0.01) | 29.67 (0.01) |
| Diabetes (%) | 44,961 (51.45) | 120,168 (26.70) | 165,129 (30.72) | 1,488 (45.04) | 8,487 (17.04) | 9,975 (18.78) | 46,449 (51.23) | 128,655 (25.74) | 175,104 (29.65) |
| History of TIA/Stroke (%) | 25,210 (28.85) | 44,222 (9.83) | 69,432 (12.92) | 815 (24.67) | 2,672 (5.36) | 3,487 (6.57) | 26,025 (28.70) | 46,894 (9.38) | 72,919 (12.35) |
| CKD (%) | 35,601 (40.74) | 48,470 (10.77) | 84,071 (15.64) | 959 (29.03) | 2,294 (4.61) | 3,253 (6.12) | 36,560 (40.31) | 50,764 (10.16) | 87,324 (14.79) |
| Hypertension (%) | 84,389 (96.57) | 312,637 (69.46) | 397,026 (73.87) | 3,028 (91.65) | 23,311 (46.90) | 26,339 (49.59) | 87,417 (96.39) | 335,948 (67.21) | 423,365 (71.69) |
| Hyperlipidemia (%) | 79,961 (91.50) | 323,071 (71.78) | 403,032 (74.99) | 2,840 (85.96) | 26,336 (52.88) | 29,176 (54.93) | 82,801 (91.30) | 349,407 (69.90) | 432, 208 (73.19) |
| CAD (%) | 66,624 (76.24) | 100,803 (22.40) | 167,427 (31.15) | 1,935 (58.57) | 3,846 (7.72) | 5,781 (10.88) | 68,559 (75.59) | 104,649 (20.94) | 173,208 (29.33) |
| Atrial Fibrillation (%) | 40,865 (46.76) | 41,807 (9.29) | 82,672 (15.38) | 930 (28.15) | 1,210 (2.43) | 2,140 (4.03) | 41,795 (46.08) | 43,017 (8.61) | 84,812 (14.36) |

Values are presented as mean (standard error) for continuous variables and n (%) for categorical variables.
HF, heart failure; HFrEF, heart failure with reduced ejection fraction; HFpEF, heart failure with preserved ejection fraction; BMI, body mass index; TIA, transient ischemic attack; CKD, chronic kidney disease; CAD, coronary artery disease.

### X-Chromosome-Wide Significant Loci

In the multi-ancestry XWAS meta-analysis, among males, five loci were associated with all-cause HF, including *FTSJ1, SPIN3, DLG3, BRWD3*, and *CHRDL1*; for HFrEF, five loci reached significance in *SPIN3, BRWD3, CHRDL1, FHL1* and *GABRA3*; for HFpEF, only one locus near *DLG3* was significant. In sex-combined analyses, six loci were associated with all-cause HF, including *FTSJ1, PAGE3, SPIN3, BRWD3, CHRDL1*, and *KIAA1210*, and four loci were associated with HFrEF, including *SPIN3, CHRDL1, FHL1*, and *GABRA3*. The *FTSJ1* locus was specific to all-cause HF, whereas FHL1 and GABRA3 were HFrEF-specific; *PAGE3* and *KIAA1210* locus were specific to sex-combined all-cause HF. The *GABRA3* signal were observed in both male-specific and sex-combined HFrEF analysis with different lead SNPs (rs17320283 and rs4828685, respectively), which are in the same LD block (r^2^ >0.8 across all populations). No loci reached significance in female-specific analyses, **Table 2, Figures 2 and S1**. Among the identified loci, variants near *PAGE3* and *SPIN3* exhibited heterogeneity across ancestries. The lead SNP at *SPIN3* locus demonstrated significant heterogeneity in male-specific and sex-combined HFrEF meta-analyses (P=0.02 and 6.82E-04), whereas this heterogeneity was not statistically significant for all-cause HF (P=0.06 and 0.08), suggesting greater variability in effect allele frequencies or effect size within the HFrEF subtype. For replication in the UKB, the signal near *BRWD3* replicated in the male replication analysis and the sex-combined replication analysis (p=0.019 and 0.039, respectively, same direction of effect). rs2281868 near *DLG3* locus showed suggestive evidence of replication (p =0.052, same direction of effect), just above the nominal significance threshold of p<0.05, **Table 2**.

**Table 2.**
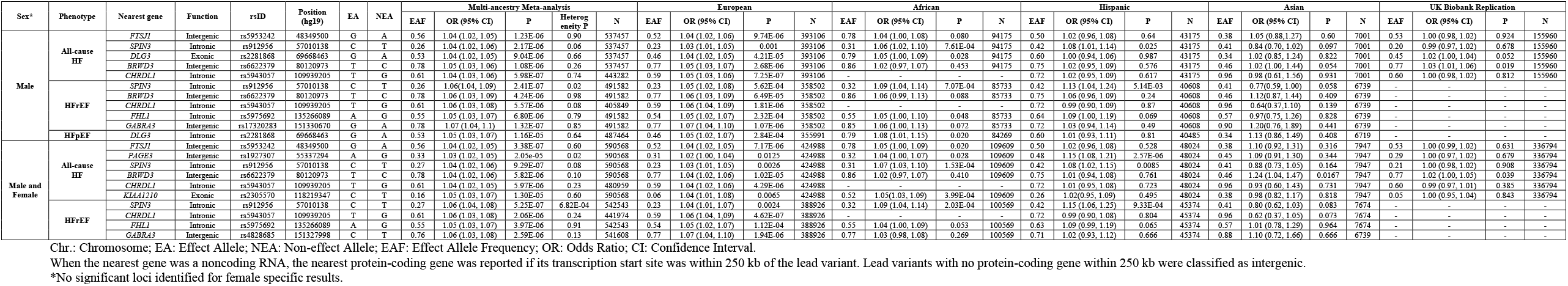
Significant Loci for All-cause HF, HFrEF and HFpEF from the Multi-ancestry XWAS Meta-analysis.

**Figure 2.**
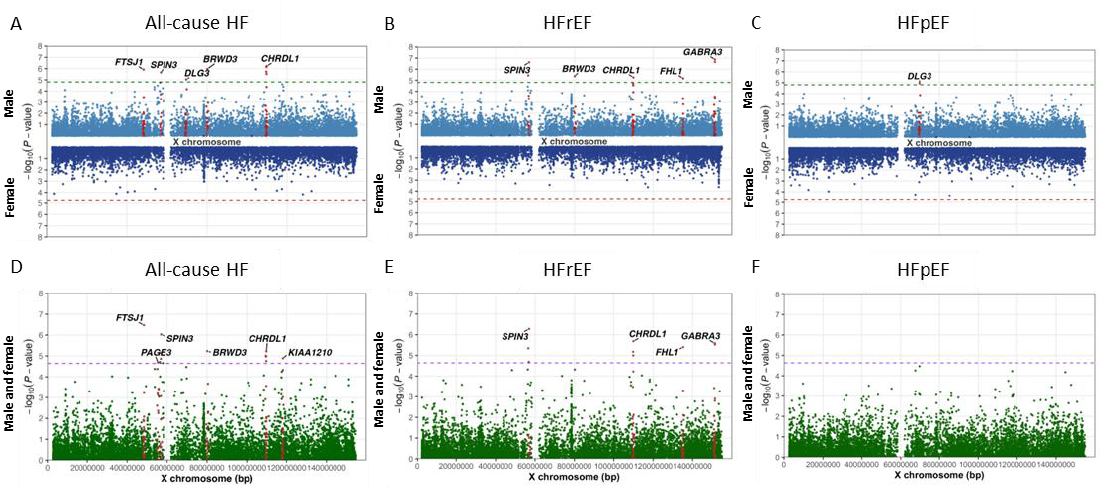
Manhattan Plots of the Multi-Ancestry X-Chromosome Association Studies of All-cause HF, HFrEF and HFpEF. The y-axis shows −log_10_ (P-value), and the x-axis indicates position along the X chromosome. The Bonferroni-corrected significance threshold, calculated as 0.05 divided by the number of effective tests, is indicated by dashed line.

In ancestry-stratified XWAS, we observed both shared and ancestry-specific associations across HF phenotypes. In European ancestry, significant signals for all-cause HF were detected in *FTSJ1, BRWD3* loci (identified lead SNPs to multi-ancestry meta-analysis) and RTL9 locus in male and sex-combined analyses. The signal at the RTL9 locus (lead SNP rs2073787) was detected in high LD (r2 >0.7 in Europeans) with the meta-analysis lead SNP at CHRDL1 (rs5943057). For HFrEF, the associated loci comprised *GABRA3* (matching the sex-combined HFrEF meta-analysis), and *RTL9* locus (lead SNP rs12013156; r2>0.8 in Europeans with rs5943057 at *CHRDL1* locus). In African ancestry, we identified a locus near *NDP* associated with male specific all-cause HF and HFpEF, a female-specific HFrEF association in *CHM*, and a sex-combined HFpEF association near *WDR44*. In Hispanic ancestry, *PAGE3* locus (identified in multi-ancestry all-cause HF) demonstrated consistent association in both male-specific and sex-combined analyses of all-cause HF and HFrEF. Additional all-cause HF associations were observed in sex-combined analyses near *DDX53, PCDH11X* locus, and HFrEF signals including *PCDH11X, PHF8*, and *GRP50* loci; Hispanic female-specific associations were also detected for HFrEF and HFpEF, though based on smaller sample sizes. In Asian ancestry, a sex-combined HF association was found at *CDR1* locus (lead SNP rs6636093). The same *CDR1* locus was also identified in Hispanic sex-combined HFpEF with a different lead SNP (rs7882637), representing an independent signal (r2<0.1). Replication in UK Biobank was observed for the European-ancestry signal near *BRWD3*, **Table 3 and Figures S2-S6**.

**Table 3.**
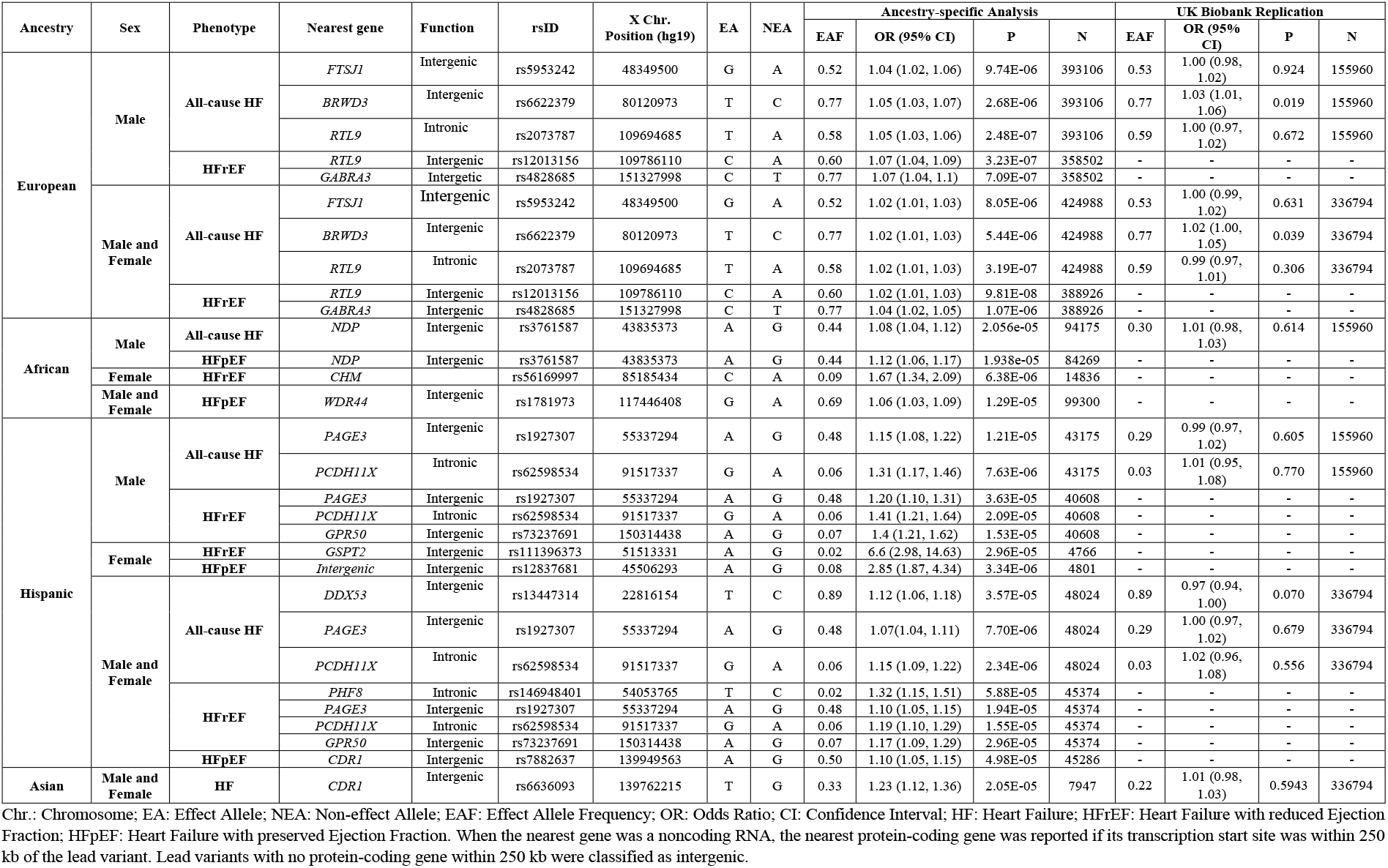
Significant Loci for All-cause HF, HFrEF and HFpEF from the Ancestry-specific XWAS.

### Gene-based Association Test and Cell Type Enrichment Analysis

Gene-based testing using MAGMA supported several loci identified in our single-variant analyses, **Tables S3-S4**. In the multi-ancestry meta-analysis, six genes achieved statistical significance. *SPIN3* was associated with both male-specific and sex-combined analyses of all-cause HF and HFrEF. *CHRDL1* and *FTSJ1* showed associations with all-cause HF whereas *GABRA3* was specially linked to HFrEF. *TBC1D25* exhibited male-specific significance while *DOCK11* was significant only in the sex-combined analyses. Ancestry-specific gene-based results further support associations in *FTSJ1* and *RTL9* in individuals of European ancestry. Surprisingly, *MAOB* demonstrated a significant association in the African ancestry group.

To explore the cellular contexts underlying HF genetic associations, we performed cell-type specificity analysis using MAGMA gene-property tests in FUMA, integrating scRNA-seq data from heart (left ventricle), kidney, liver, and skeletal muscle tissues. Significant enrichment after multiple-testing correction was observed predominantly for natural killer (NK) cells and related innate/adaptive immune subsets, **Figure S7**. Within-dataset conditional analyses confirmed that associations with mucosal invariant T cells and NK subsets remained independent after controlling for correlated cell types within each reference. Low-to-moderate cross dataset proportional significance values in cross-dataset conditional analyses suggested partial overlaps among immune-related cell-type enrichments, consistent with shared transcriptional programs across lymphoid subsets. These findings implicated systemic immune dysregulation, particularly involving NK-cell mediated processes, in HF genetic risk.

### Tissue Expression Analysis and cis-eQTL

In tissue expression analyses using data from Genotype-Tissue Expression (GTEx) v8 dataset, significant cardiovascular-related enrichment was observed for male HFrEF in the aorta, **Tables S5-S6**. To identify the potentially causal genes, we examined cis-eQTL associations for the identified lead SNPs from the risk loci in the GTEx v8, focusing on heart relevant tissues, **Tables S7-S8**. Six lead SNPs from nine identified risk loci, associated with HF or its subtypes across all meta-analyses, show at least one significant eQTL(FDR ≤ 0.05). Specifically, rs912956 was associated with *SPIN3, SPIN2B, KLF8*, and *FAAH2*, across all nine heart-relevant tissues. rs5943057 showed strong associations with *CHRDL1* expression in tibial artery (normalized effect size: NES=-0.33, p=1.22E-48), skeletal muscle (NES=-0.16, p=2.36E-17), aorta (NES=-0.11, p=1.14E-05) and left ventricle (NES=-0.12, p=6.61E-05), and with *RTL9* in skeletal muscle (NES=0.17, p=1.87E-07) and Heart left ventricle (NES=0.15, p=4.28E-05). Four of five European-specific lead SNPs showed significant eQTL associations. rs1781973 at *WDR44* locus in African ancestry was associated with *WDR44* expression (NES=0.05, p=6.74E-5) in whole blood tissue, and rs3761587 near *NDP* locus displayed an exclusive eQTL association with *NDP* in cerebellum brain tissue. For lead SNPs in Hispanic ancestry, one of seven lead SNP (rs1927307) exhibited significant eQTL associations with *MAGEH1* and *TRO*.

### Credible Sets and Colocalization Characterize Risk Loci

Credible set fine mapping highlighted a subset of loci with high resolution, defined as fewer than 20 variants in the posterior probability 0.90 or 0.95 credible sets. In the multi ancestry meta-analysis, high resolution credible sets were observed at *SPIN3* and *BRWD3* in males, and at *FTSJ1, BRWD3*, and *FHL1* in sex combined or male HFrEF analyses, **Table S9**. In ancestry specific analyses, high resolution credible sets were observed for *BRWD3* in Europeans, for the HFpEF signal in Africans, for selected Hispanic loci including *PHF8*, **Table S10**.

Colocalization analysis was performed using variants within ±250 kb of the lead SNP (rs5943057) from the male-specific and sex-combined meta-analyses of HF and HFrEF, integrated with eQTL data from GTEx. Suggestive evidence of colocalization was observed exclusively in skeletal muscle tissue, with Bayesian posterior probabilities of a shared causal variant (PPH4) ranging from 0.50 to 0.75 **Figure S8**. These findings provide moderate support that HF and HFrEF association signals share a causal regulatory variant influencing the eQTL signals for CHRDL1 and RTL9 in skeletal muscle. For other loci examined, the density of variants within the ±250 kb window was insufficient to enable reliable colocalization analysis.

### Phenome-wide Association Study

Based on the UK Biobank PheWeb (https://pheweb.org/UKB-TOPMed/), four lead SNPs of rs5943057 at *CHRDL1*, rs5975692 at *FHL1*, rs12013156 and rs2073787 near *RTL9* showed strong association (p<1e-5) with cardiovascular related traits, including coronary atherosclerosis, ischemic heart disease and hyperlipidemia, **Table S11-S12, Figures S9-S10**. These findings suggest potential pleiotropic effects of the identified risk loci on cardiometabolic traits.

## Discussion

This multi-ancestry XWAS provides the first large-scale evidence of X-linked genetic contributions to HF and its subtypes. Loci such as *SPIN3* and *CHRDL1* that showed significant associations across male-specific and sex-combined analyses for HF and HFrEF indicate shared biological mechanisms underlying these phenotypes. In contrast, *FTSJ1* and *KIAA1210* exhibited associations specific to all-cause HF, whereas *FHL1* and *GABRA3* were HFrEF-specific. Only *DLG3* locus emerged in HFpEF. Ancestry-stratified analyses further revealed conserved signals (e.g., *FTSJ1, BRWD3, RTL9* in Europeans; *PAGE3* in Hispanics), as well as population-specific loci, such as *NDP* and *WDR44* (African ancestry), *PCD11X/PHF8/GPR50* (Hispanic ancestry), and *CDR1* (Asian ancestry). These findings extend prior large-scale autosomal HF GWAS efforts by highlighting X-chromosomal contributions.

Loci identified by our analyses mapped to genes with biologically plausible links to cardiac structure, remodeling, and heart failure susceptibility. The *CHRDL1*–*RTL9* locus is particularly notable, as *CHRDL1* encodes a secreted regulator of bone morphogenetic protein (BMP) signaling, a pathway implicated in cardiac hypertrophy, fibrosis, and myocardial infarction, supporting a role for this region in adverse remodeling and HF risk[33, 34]. Notably, *RTL9* has been reported to be significantly associated with a cardiovascular condition, i.e., sporadic thoracic aortic dissection in an X-chromosome–wide association study [35]. Suggestive colocalization, robust cis-eQTL evidence in heart-relevant tissues, and pleiotropic associations with cardiometabolic traits, reinforce the likelihood that regulatory mechanisms at *CHRDL1*– *RTL9* locus contribute to heart failure susceptibility. BRWD3 has been linked to adiposity, a well-established risk factor for heart failure [36]. FHL1 is associated with X-linked myopathy with cardiac involvement and cardiomyopathy phenotypes [37]; it functions as a sarcomeric mechanotransducer at the titin N2B region, sensing biomechanical overload and activating stress-responsive signaling pathways that promote pathological cardiac hypertrophy/remodeling [38]. PHF8 is postulated to act as an endogenous protective factor against pathological remodeling, hypertrophy, fibrosis, cardiomyocyte apoptosis, and oxidative stress in heart failure models [39, 40]. Novel loci such as *SPIN3* (a chromatin reader involved in epigenetic regulation) and *FTSJ1* (a tRNA methyltransferase implicated in translational control under stress) suggest roles in epigenetic and post-translational control of cardiac stress responses[41]. *WDR44* and *NDP* regulate primary ciliogenesis and Wnt signaling during development [42, 43]. Beyond locus-specific mechanisms, cell-type specificity analyses implicated systemic immune dysregulation, particularly NK-cell–mediated processes, in HF genetic risk. This finding aligns with emerging evidence that NK cells modulate cardiac inflammation, fibrosis, and immune homeostasis during cardiac injury and remodeling [44, 45]. Collectively, the identified X-chromosomal loci converge on clinically relevant biological pathways, including BMP signaling, biomechanical stress response, epigenetic or transcriptional control, ciliogenesis/Wnt signaling and immune regulation, that likely contribute to heart failure susceptibility and subtype progression.

In contrast to prior large-scale efforts, including a comprehensive X-chromosome-wide meta-analysis of coronary artery disease involving over 101,000 individuals that identified no genome-wide significant associations with X-linked variants[46], our study uncovered several robust X-chromosomal gene associations relevant to heart failure subtypes. These findings underscore the value of sex-stratified and ancestry-stratified analyses, as well as the newly developed X-Chromosome-Wide significance threshold approach (detailed in the Methods). Notably, these discoveries were achieved using only raw genotyped variants (∼11,000 after quality control) from the Million Veteran Program, without imputation to denser reference panels. This strategy prioritizes high-confidence genotype calls over imputation. The exceptional scale and ancestry diversity of the MVP cohort (approximately 28% non-European ancestry, including ∼18% African ancestry) provided sufficient statistical power to detect novel associations among directly genotyped variants. The large-scale and diverse ancestry composition of MVP cohort also partially explains the modest or limited replication observed in UK Biobank, where BRWD3 replicated nominally in male-specific and sex-combined HF analyses. Key cohort differences include MVP’s male predominance, multi-ancestry diversity, veteran-based ascertainment with high comorbidity burden, versus UK biobank’s more sex-balanced, predominantly European ancestry, which likely reduced replication power.

Despite the large sample size and quality control steps to address methodological issues with XWAS, our study has some limitations. The predominance of discoveries in males and sex-combined models likely reflects both biology and higher statistical power in the MVP, while the lack of female-specific loci should be interpreted cautiously given smaller sample size. Similarly, the relative paucity of HFpEF signals is consistent with the broader GWAS literature in which HFpEF has historically yielded fewer loci than HFrEF, likely reflecting greater phenotypic heterogeneity [8, 47]. However, differences between HFrEF and HFpEF results must be interpreted with caution in this study. The MVP cohort’s strong male bias, combined with focus on X-chromosome variation (where males are hemizygous), and the well-established sex differences in HF subtype prevalence and pathophysiology, creates a complex interpretive landscape and limits strong conclusions regarding subtype specific X-linked genetic contributions. Additionally, while the male-predominant cohort may have limited discovery in females, it offers a potential advantage in terms of associations discovered in males since genomic imprinting and allelic dosage issues do not affect variant association in males.

Future work should expand X-chromosome discovery beyond genotyped markers by leveraging sequencing across multiple large biobanks with improved representation of women and non-European ancestry groups, thereby increasing power to detect female-specific and ancestry-specific effects and enabling more complete assessment of common and rare X-linked variation. In parallel, the highest-priority loci emerging from fine-mapping and colocalization should be advanced to experimental validation of regulatory elements and gene perturbation in relevant human cell systems, and to clinical validation in independent cohorts with harmonized HF subtyping and longitudinal outcomes, to clarify whether X-linked regulatory mechanisms can be translated into sex-informed prediction, prevention, and therapeutic targeting for HF and its subtypes.

## Conclusions

Our multi-ancestry, sex-stratified, XWAS in a carefully phenotyped, large national biobank identified X-linked genetic contributions to HF and its subtypes, highlighting the value of including the X chromosome to better understand genetic risk of HF.

## Acknowledgement

This research is based on data from the Million Veteran Program, Office of Research and Development, Veterans Health Administration, and was supported by MVP000, MVP037 (Dr. Phillips, Dr. Sun), MVP065 (Dr. Joseph, Dr. Jacobson) as well as awards CX001737, BX005831, BX004821. This publication does not represent the views of the Department of Veteran Affairs or the United States Government. This research has also been supported in part by National Institutes of Health (NIH) grant P01 HL154996.

The research was conducted using data from the UK Biobank Resource under application number 34031. The UK Biobank will make the data available to all bona fide researchers for all types of health-related research that is in the public interest, without preferential or exclusive access for any persons. All researchers will be subject to the same application process and approval criteria as specified by UK Biobank. For more details on the access procedure, see the UK Biobank website: www.ukbiobank.ac.uk.

## Data Availability

Due to US Department of Veterans Affairs (VA) regulations and our ethics agreements, the analytic datasets used for this study are not permitted to leave the Million Veteran Program (MVP) research environment and VA firewall. This limitation is consistent with other MVP studies based on VA data. However, the MVP data are made available to researchers with an approved VA and MVP study protocol. The full summary level genome-wide association analyses results will be available through dbGaP with accession number phs001672.

MVP Program Office

VA Million Veteran Program

Core Acknowledgements for Publications October 2025

- Sumitra Muralidhar, Ph.D., Program Director US Department of Veterans Affairs, 810 Vermont Avenue NW, Washington, DC 20420
- Jennifer Moser, Ph.D., Associate Director, Scientific Programs US Department of Veterans Affairs, 810 Vermont Avenue NW, Washington, DC 20420
- Jennifer E. Deen, B.S., Associate Director, Cohort & Public Relations US Department of Veterans Affairs, 810 Vermont Avenue NW, Washington, DC 20420

MVP Steering Committee

- Co-Chair: Philip S. Tsao, Ph.D. VA Palo Alto Health Care System, 3801 Miranda Avenue, Palo Alto, CA 94304
- Co-Chair: Sumitra Muralidhar, Ph.D. US Department of Veterans Affairs, 810 Vermont Avenue NW, Washington, DC 20420
- J. Michael Gaziano, M.D., M.P.H. VA Boston Healthcare System, 150 S. Huntington Avenue, Boston, MA 02130
- Adriana Hung, M.D., M.P.H., VA Tennessee Valley Healthcare System, 1310 24th Avenue, South Nashville, TN 37212
- Dave Oslin, M.D. Philadelphia VA Medical Center, 3900 Woodland Avenue, Philadelphia, PA 19104
- Deepak Voora, M.D. Durham VA Medical Center, 508 Fulton Street, Durham, NC 27705

MVP Co-Principal Investigators

- J. Michael Gaziano, M.D., M.P.H. VA Boston Healthcare System, 150 S. Huntington Avenue, Boston, MA 02130
- Philip S. Tsao, Ph.D. VA Palo Alto Health Care System, 3801 Miranda Avenue, Palo Alto, CA 94304

MVP Core Operations

- Jessica V. Brewer, M.P.H., Director, MVP Cohort Operations VA Boston Healthcare System, 150 S. Huntington Avenue, Boston, MA 02130
- Mary T. Brophy M.D., M.P.H., Director, VA Central Biorepository VA Boston Healthcare System, 150 S. Huntington Avenue, Boston, MA 02130
- Kelly Cho, M.P.H, Ph.D., Director, MVP Phenomics VA Boston Healthcare System, 150 S. Huntington Avenue, Boston, MA 02130
- Lori Churby, B.S., Director, MVP Regulatory Affairs VA Palo Alto Health Care System, 3801 Miranda Avenue, Palo Alto, CA 94304
- Jacob T. Kean, Ph.D., Acting Director, VA Informatics and Computing Infrastructure (VINCI) VA Salt Lake City Health Care System, 500 Foothill Drive, Salt Lake City, UT 84148
- Saiju Pyarajan Ph.D., Director, Data and Computational Sciences VA Boston Healthcare System, 150 S. Huntington Avenue, Boston, MA 02130
- Robert Ringer, Pharm.D., Director, VA Albuquerque Central Biorepository New Mexico VA Health Care System, 1501 San Pedro Drive SE, Albuquerque, NM 87108
- Luis E. Selva, Ph.D., Director, MVP Biorepository Coordination VA Boston Healthcare System, 150 S. Huntington Avenue, Boston, MA 02130
- Shahpoor (Alex) Shayan, M.S., Director, MVP PRE Informatics VA Boston Healthcare System, 150 S. Huntington Avenue, Boston, MA 02130
- Brady Stephens, M.S., Principal Investigator, MVP Information Center Canandaigua VA Medical Center, 400 Fort Hill Avenue, Canandaigua, NY 14424
- Stacey B. Whitbourne, Ph.D., Director, MVP Cohort Development and Management VA Boston Healthcare System, 150 S. Huntington Avenue, Boston, MA 02130

